# Multimodality Integration of Neural Social Activation and Social and Language Scores Reveals Three Replicable Profound and Milder Autism Subtypes With Divergent Clinical Outcomes

**DOI:** 10.1101/2024.05.30.24308230

**Authors:** Vani Taluja, Sanaz Nazari, Javad Zahiri, Lana Garmire, Karen Pierce, Yaqiong Xiao, Eric Courchesne

## Abstract

Social affective and communication symptoms stand at the center of autism, and usually become apparent within the first 1-3 years of life. Symptom severity differs widely across toddlers and clinical outcomes, ranging from near-neurotypical to poor. The biological bases of this early and wide symptom diversity are largely unknown. While more than two dozen studies have attempted to subgroup early-age clinical heterogeneity, most studies fail to rigorously validate discovered subtypes using multiple methods, and none linked observed clinical subtypes with underlying functional neural signatures. Using a well-established approach for precision medicine patient subtyping (Similarity Network Fusion) and multiple rigorous validation methods, we integrated thoroughly replicated measures of social neurofunctional activation and social and language ability in 137 toddlers at early ages. Results identified three distinct social neural-clinical ASD subtypes, validated using multiple methods. One subtype was consistent with a ‘profound’ autism profile with negligible social neural activation, severe social and language symptoms, low social interest, and little clinical improvement. Another ASD subtype had a contrasting pattern with only mildly reduced social neural activity, near neurotypical social and language abilities, and substantial age-related clinical improvement. One principal implication of these results is that the “spectrum” of ASD heterogeneity is not truly a continuous spectrum from the neurobiological and clinical perspective. The profound autism subtype is the neurofunctional, clinical and developmental opposite of the mild ASD subtype, suggesting different etiological mechanisms. A second implication is that neurobiological and clinical subtype differences highlight the need to develop subtype-specific treatments, particularly for the profound subtype. Third, treatment studies with an undetermined mix of subtypes could fail or succeed based on how many patients from each subtype are included in the mix.

## INTRODUCTION

Social affective and communication symptoms appear in the first years of life in autism spectrum disorder (ASD) and give rise to pervasive developmental challenges, yet to differing degrees. Some ASD toddlers have outcomes considered good or “high” ability while many do not, but neurofunctional reasons for differential outcomes are neither known nor predictable at the youngest ages. Children with the most severe symptoms, or so-called “profound” autism^1,2^, are at the greatest risk for a poor outcome, including life-long struggle with ASD symptoms. However, due to low levels of their inclusion in imaging research studies, particularly in functional brain imaging studies that require holding still, little is known about the neural basis of social and language symptoms in this important group of autistic toddlers. Because impairments in social perception and reactivity are early and specific signs of ASD, there is a particular necessity to uncover the exact neurobiological differences that underlie different ASD clinical subgroups.

Over 25 studies of ASD toddlers used exclusively clinical diagnostic, psychometric and/or behavioral measures to cluster patients, commonly finding 2 to 4 clusters^3^. All were weakly validated by only one or two methods (e.g., cross-method replication) and some used no validation at all^3^. None of these 25 studies incorporated into the clustering both neurobiological measures relevant to social affective and communication symptoms and clinical scores. Thus, these studies provide no insight into the neural bases of clinical social subgroups, and only weak understanding of clinical subgroups since rigorous cluster validations were absent. Further, many clustering designs used only a single clinical measure, such as the ADOS, which limits the insights generated to a single dimension.

To gain insight into the neural bases of clinical social subgroups, one strategy utilizes experimenter-defined clinical stratification in conjunction with neurobiological measures relevant to social affective and communication symptoms^4^. To assess social neural functional variability across all ability levels at the age of first social symptoms, in our studies we leveraged sleep functional imaging and passive listening to social affect speech. In this design, we stratified ASD toddlers into two groups based on language ability and found that ASD toddlers with good language outcomes had strong temporal cortex activation to social speech, and that those with poor outcomes have much weaker activation. This stratification design showed that underlying social symptoms in autism is temporal cortex dysfunction in response to social stimuli.

A different strategy is to use unbiased, unsupervised precision medicine methods such as Similarity Network Fusion (SNF) to objectively discover patient subtypes. SNF is a well-established method that integrates any type of multimodality data (e.g., clinical, biological) to reveal distinct multimodal subtypes among patient and/or control groups, wherein the multimodality profile of patients in a cluster are maximally similar to each other and maximally different from those of patients in other clusters^5^. Using SNF, we integrated measures of fMRI social activation in temporal and frontal cortices with clinical measures in ASD and typically developing (TD) toddlers^6^. We showed temporal cortex measures of fMRI activation to social stimuli are positively correlated with clinical measures in ASD and TD toddlers, specifically social and language abilities. There were three distinct social neural-clinical subtypes among ASD subjects, one with slightly reduced temporal cortex activation and high clinical ability, another with both moderately reduced activation and ability, and a third with weak social activation and low social ability. To our knowledge, this was the first unsupervised precision social brain-behavior study in ASD^6^.

This discovery of dysfunctional social activation of temporal cortex in ASD at the earliest ages has major importance because temporal cortex is a hub of social information processing in the typical brain^7–14^. Moreover, ASD temporal cortex anatomical enlargement is a feature predictive of early age language outcome^15^. ASD temporal cortex is also the only cortical region that is persistently enlarged across ASD ages from toddlers to adults^15–17^. Remarkably, ASD temporal cortex along with visual regions are the top two cortical hot spots for differentially expressed (DE) genes, with 2,733 and 3,264 DE genes, respectively, both of which are far more than all other cortical regions (e.g., 409 DE genes in frontal, 0 DE genes in fusiform)^18^.

Knowledge of ASD social neural and clinical subtypes is vital for advances towards understanding subtype-specific etiology, neurobiology, and especially treatment in this heterogenous disorder. As such, recently identified social neural-clinical subtypes must be rigorously validated. Here, using a sample that is more than three times the size of our initial study^6^, we rigorously validate social neural-clinical subtypes of ASD using data-driven SNF methods^5^ with a large dataset of deeply phenotyped ASD, typically developing, and neurodevelopmentally delayed subjects. We leverage state of the art bioinformatic methodology and multimodal SNF analyses (behavioural, cognitive, and neurofunctional) to reveal clinically meaningful toddler-age social-neural subtypes of ASD. With this approach, we identified ASD neural-clinical subtypes that have categorically opposite clinical developmental trajectories: profound autism and mild autism. Then to contextualize ASD subtypes in relation to the broader non-ASD toddler population, we mapped them with comparable data from a range of typical and developmentally delayed subjects again using unsupervised SNF which revealed a potential “optimal” outcome subtype within the mild ASD subtype.

## METHODS

### Participants

This study was approved by the University of California, San Diego Institutional Review Board. Informed consent was obtained from parents or guardians of toddlers. Participants were recruited via the *GET SET Early* Approach^19^. Sleep fMRI and social eye tracking data were collected from a total of 139 toddlers (2 were excluded for head motion during fMRI scan). Of the remaining 137 toddlers, 81 were diagnosed with ASD and 56 were non-ASD control toddlers, including 33 with typical development (TD) and 23 with a delay (7 with ASD features, 3 with language delay (LD), and 13 with global developmental or other delays). Of the 20 non-LD delayed toddlers, 6 had features of “late talkers”^20^, namely low clinical scores at intake driven by low expressive language (mean EL score: 66.09), followed by improvements on the order of 1-4 standard deviations (mean EL score: 95.94) to near neurotypical clinical profiles about a year later.

Initial clinical data were collected around 1 – 3 years of age, including the Autism Diagnostic Observation Schedule (ADOS-2; Module T, 1, or 2)^21^, the Mullen Scales of Early Learning (MSEL)^22^, and the Vineland Adaptive Behavior Scales (VABS)^23^. The same clinical data were collected again from all subjects longitudinally about a year after intake, around 2 – 4 years of age.

Clinical, behavioral, and neuroimaging data were collected between 2008 and 2014; part of the neuroimaging data were reported in previous studies^4,24,25^ and eye-tracking data were included in previous publications.^26,27^ For demographic characteristics and clinical measures at intake and outcome, see Supplementary Table S1. SNF analyses of these data in the present work are reported here for the first time.

### Affective language paradigm

The affective language paradigm was identical to our previous studies^4,6,24,25,28^, and included three types of stimuli: age-matched forward speech, age-advanced forward speech, and backward speech. Stimuli were presented in a block design, with each stimulus presented 20 sec and followed by 20 sec of rest (9 blocks, 6 min 24 sec). As activation to forward and backward speech stimuli did not differ in previous studies using the identical paradigm^4,6,24,25^ nor in this work, we combined forward and backward speech stimuli in the analysis, with speech versus rest as the main contrast of interest.

### Imaging data acquisition and analyses

All fMRI data were collected using a 1.5 Tesla GE MRI scanner (GE High-Definition 1.5 T twin-speed EXCITE scanner) from all toddlers during natural sleep at the University of California, San Diego. Functional images were acquired with echoplanar imaging (TE = 35ms; TR = 2500ms; flip angle = 90°; matrix size = 64 × 64; resolution = 4 × 4 mm; slice thickness = 4 mm; FOV = 256 mm; 31 slices; 154 volumes). Structural images were acquired using a T1-weighted MPRAGE sequence (FOV = 228 mm; matrix size = 256 × 256; resolution = 0.89 × 0.89 mm; slice thickness = 1.5 mm; 128 slices).

Functional imaging data were preprocessed using AFNI^29^, including motion correction, normalization to Talairach space, and smoothing with a 8 mm FWHM Gaussian kernel. The first-level and second-level whole-brain activation analyses were modeled with the general linear model in SPM8 (http://www.fil.ion.ucl.ac.uk/spm8/). Events in first-level models were modeled with the canonical hemodynamic response function and its temporal derivative, with motion parameters as covariates of no interest. High-pass temporal filtering was applied with a cutoff of 0.0078 Hz (1/128 s) to remove low-frequency drift in the time series.

Analysis of head motion via framewise displacement (FD) showed two subjects had considerable head motion (mean FD > 0.5) and were excluded in the sample. For the remaining subjects, head motion was minimal (mean FD < 0.25) for nearly all subjects across diagnostic groups (ASD, mean = 0.08 mm, s.d. = 0.09; TD: mean = 0.07 mm, s.d. = 0.03; ASD Features, mean = 0.06 mm, s.d. = 0.04; DD/Other, mean = 0.1 mm, s.d. = 0.07; LD: mean = 0.12 mm, s.d. = 0.11), and groups did not differ in mean FD (*F*(4, 169) = 0.93, *p* = 0.45). Percent signal change (speech versus rest) was extracted based on beta maps from the first-level models for frontal and temporal language regions using Neurosynth ROIs (with the term ‘language’) as those used in previous studies^4,6,24^.

### SNF Analyses

We used early-age fMRI brain activation to speech once with outcome clinical measures and again with intake clinical measures to design two separate outcome and intake SNF analyses^5^ (‘SNF’ function in the ‘*SNFtool*’ package). Then, we used a spectral clustering algorithm (‘spectralClustering’ function in the ‘*SNFtool*’ package) to detect clusters of the similarity network with the ASD sample. In these analyses, we included fMRI activation data (four variables: bilateral frontal and temporal ROI percent signal change) as well as outcome or intake clinical data respectively (three variables: MSEL standardized age equivalent scores for receptive and expressive language subscales, and VABS adaptive behavior composite) for all 81 ASD subjects. We determined three to be the optimal number of clusters using various indices and clustering methods in several R packages, including *NbClust*^30^, *mclust*^31^, and *SNFtool*.

For the whole sample (n = 137) including 81 ASD subjects and 56 non-ASD controls, we ran the SNF and clustering analysis using the same approach as for the ASD-only sample. We then analyzed how subjects in each of the ASD Outcome clusters, as identified by SNF, mapped onto each of the Mixed clusters with the whole sample of ASD and non-ASD subjects. Correlation analyses were conducted among the four fMRI activation scores and MSEL receptive and expressive standardized age equivalent scores for both Outcome and Mixed SNFs.

### Validation strategies for ASD Outcome subtypes and Mixed subtypes

We used five methods^3^ to validate obtained clusters from Outcome SNF and six to validate Mixed SNF. First, we tested cluster separation based on each variable included in SNF to determine if any factors were driving cluster assignment, and to quantify the degree of certainty with which patients are assigned to specific clusters. Second, to establish external validity, cluster separation was also tested on clinical and social attention variables not used in the SNF, including MSEL Early Learning Composite (ELC), ADOS total score, and fixation on social images during eye tracking. Third, we ran a prediction analysis with the 5-fold cross-validation approach on Outcome and Mixed SNF. Specifically, the whole sample was randomly divided into 5 equal-sized parts: one part was used as the left-out test dataset, and the rest was used as the training dataset. Then, using the label propagation method, the cluster labels were predicted for the left-out samples. This process was repeated five times, using each of the five parts as the left-out test set once. The cluster label prediction was performed using the ‘groupPredict’ function in the ‘*SNFtool*’ package in R. The 5-fold cross-validation procedure was repeated 10 times. The prediction accuracy was calculated as the percent of the accurate labels predicted by the label propagation. Fourth, we ran robustness analysis through 100 iterations with 95%, 90%, 80%, 70%, 60%, and 50% of the total sample using the SNF and clustering analysis as described above. The Normalized Mutual Information (NMI) index, calculated by the ‘calNMI’ function in ‘*SNFtool’* package, was used to measure the similarity of two clustering outcomes. For the Mixed SNF, we also used a fifth validation: independent cohort replication. This was conducted on an independent cohort of ASD and non-ASD toddlers from Xiao et. al^6^. The same SNF design with identical clinical and fMRI measures were used to test Mixed subtype replication.

### Analyses of ASD subtype differences in language and adaptive behavior developmental trajectories

To test for ASD subtype-specific differences in trajectories of longitudinal clinical language and adaptive behavior abilities, we tested whether longitudinal changes between each child’s intake and outcome clinical scores (the latter of which were used in SNF and best estimated the child’s relatively ultimate phenotype) differed by subtype cluster.

## RESULTS

### <u>Result 1:</u> Three distinct subtypes based on SNF integration of three clinical measures and fMRI response to social speech at early ages in ASD

First, we tested the hypothesis, based on our previous paper^6^, that greater fMRI activation to social speech in temporal cortex is correlated with greater receptive and expressive language ability in ASD toddlers, as is illustrated in Figure 1A. In our 81 ASD toddlers, right temporal cortex activation was significantly correlated with receptive (r=0.26, p=0.021) and expressive language (r=0.24, p=0.028), as was frontal cortex activation (RL: r=0.33, p=0.003; EL: r=0.26, p=0.017). Left temporal cortex activation was significantly correlated with receptive language (r=0.27, p=0.016) and showed a trending correlation with expressive language (r=0.21, p=0.066, see Supplementary Table S2).

**Figure 1.**
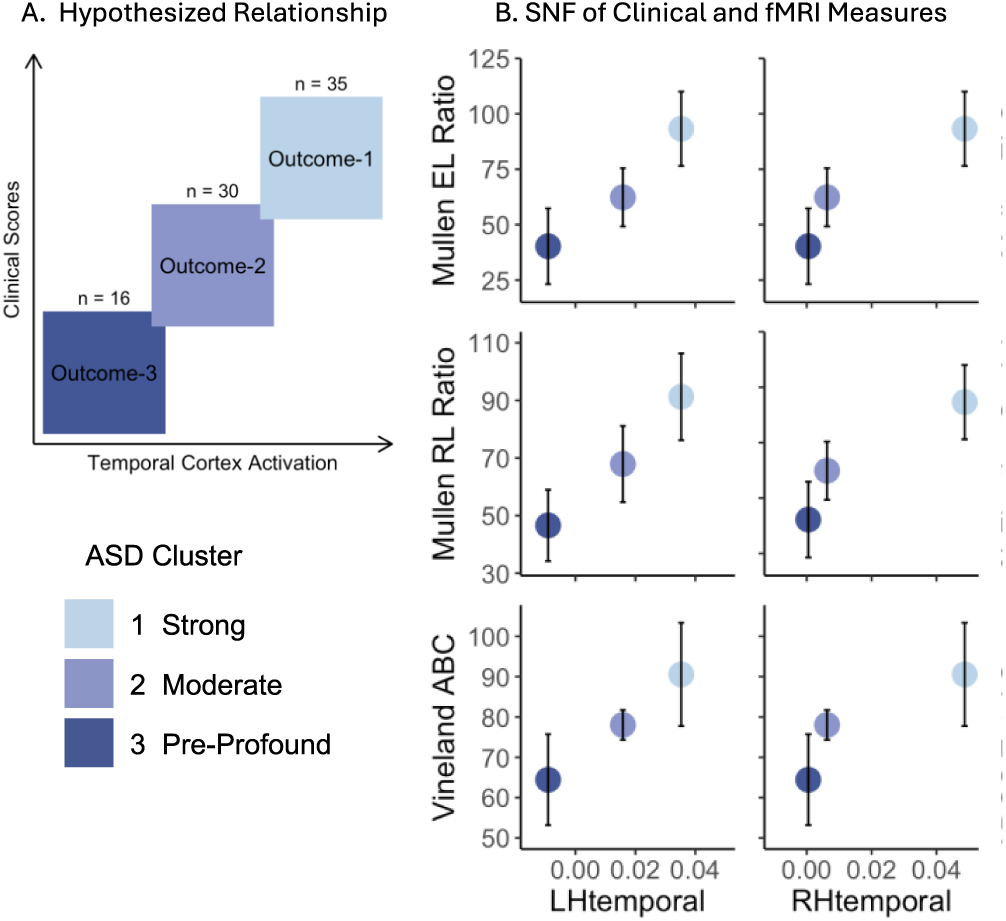
ASD Outcome SNF Clustering. Relationship between fMRI activation in temporal cortex and various clinical ability scores in ASD toddlers. (**A**) Schematic representation of hypothesized relationship. (**B**) Observed left and right hemisphere percent signal change in response to speech compared to clinical measures (expressive language, receptive language, and adaptive behavior), all of which were used in SNF clustering. See Tables 2 and 3 for statistical differences across cluster separation, all together and pairwise.

Next, leveraging clinical scores at outcome and temporal cortex fMRI activation to social speech, SNF identified three ASD Outcome clusters delineating distinct neural and clinical subtypes as shown in Figure 1B and Table 1. The fMRI activation in temporal cortex and clinical ability scores averaged for each cluster support the hypothesis illustrated in Figure 1A and are consistent with the significant group level correlations mentioned above (also see Supplementary Table 2). ASD Outcome cluster 1 subjects (n=35) showed relatively high adaptive behavior and language scores, as well as near neurotypical brain activation in response to social speech. In contrast, ASD Outcome cluster 3 subjects were the exact opposite, with social and language scores consistently more than 2 standard deviations below average, and markedly low brain activation in response to speech (Table 1, Figure 1B). Clinical scores for cluster 3 subjects are consistent with clinical characteristics of profound ASD: namely, highly impacted language (e.g., −3SD below average mean), severe social symptoms, and very low adaptive behavior abilities^1^ (Table 1). Subjects in ASD Outcome cluster 2 (n=30) fell in the middle and had clinical scores 1-2 standard deviations below neurotypical means and reduced neural responses.

**Table 1.** Average fMRI activation in temporal cortex and clinical ability scores for each ASD Outcome cluster. Cluster language means differ by more than 1SD.

|  | MSEL Receptive Language | MSEL Expressive Language | ADOS Total Score | Left Temporal Lobe Activation | Right Temporal Lobe Activation |
| --- | --- | --- | --- | --- | --- |
| ASD Outcome 1 | 91.27 ± 15.08 | 93.26 ± 16.75 | 16.86 ± 5.02 | 0.0353 ± 0.04 | 0.0487 ± 0.05 |
| ASD Outcome 2 | 67.92 ± 13.21 | 62.35 ± 13.14 | 18.23 ± 3.65 | 0.0158 ± 0.05 | 0.0063 ± 0.08 |
| ASD Outcome 3 | 46.59 ± 12.37 | 40.26 ± 17.09 | 21.88 ± 3.98 | -0.0091 ± 0.04 | 0.0005 ± 0.05 |

#### Validation of ASD Outcome Clusters

Review of 156 ASD clinical clustering studies showed most reported clusters were weakly validated by only 0 to 2 methods and none had robust validation^3^. Here, we used multiple validation approaches. First, we tested cluster separation using omnibus ANOVA and multiple pairwise comparisons, as shown in Tables 2 and 3. There was significant separation between all three clusters across all clinical variables in the SNF. For fMRI variables, such as activations in the right frontal and left temporal cortices, the low and high clusters were separated; for right temporal cortex activation, both low and medium clusters were separated from the high ability cluster. We could not observe any separation of clusters for left frontal cortex activation. Second, clinical and social attention variables not used in the SNF, including MSEL overall IQ, ADOS total score, and fixation on social images during eye tracking, were used to examine external validation. The results showed significant cluster separation for MSEL ELC and separation of medium and high ability clusters from the low cluster for ADOS total score, but no cluster separation for the eye tracking score (see Table 2 and 3). As illustrated in Figure 2A, mean scores for each of these variables showed a similar linear profile across clusters to variables included in SNF, as illustrated when compared to temporal cortex fMRI activation.

**Table 2.** Separation Validation and External Validation in ASD Outcome SNF variables.

|  | Variable | Effect | F-ratio | DFn | DFd | GES | P-value |
| --- | --- | --- | --- | --- | --- | --- | --- |
| SNF | Left Frontal Lobe Activation | Clusters | 1.84 | 2 | 78 | - | 0.165 |
|  | Right Frontal Lobe Activation | Clusters | 5.21 | 2 | 78 | 0.12 | 0.008* |
|  | Left Temporal Lobe Activation | Clusters | 5.53 | 2 | 78 | 0.12 | 0.006* |
|  | Right Temporal Lobe Activation | Clusters | 5.09 | 2 | 78 | 0.12 | 0.008* |
|  | VABS Adaptive Behavior Composite | Clusters | 38.84 | 2 | 78 | 0.50 | 0.000* |
|  | MSEL Receptive Language | Clusters | 60.94 | 2 | 78 | 0.61 | 0.000* |
|  | MSEL Expressive Language | Clusters | 71.48 | 2 | 78 | 0.65 | 0.000* |
| External | ADOS Total Score | Clusters | 7.3 | 2 | 78 | 0.16 | 0.001* |
|  | MSEL Early Learning Composite | Clusters | 49.52 | 2 | 78 | 0.56 | 0.000* |
|  | GeoPref Social Fixation | Clusters | 1.69 | 2 | 69 | - | 0.193 |

*Note.* \* = $P < .05$ , $DFn$ = Degrees of freedom for numerator, $DFd$ = Degrees of freedom for denominator, $GES$ = Generalized Eta Squared (effect size).

**Table 3.** Separation Validation. Cluster *<u>pairwise</u>* comparisons in ASD Outcome SNF variables and external variables.

|  | Variable | P-values |  |  |
| --- | --- | --- | --- | --- |
|  | Outcome Cluster | 1 – 2 | 2 – 3 | 1 – 3 |
| SNF | Left Frontal Lobe Activation | 0.893 | 0.158 | 0.158 |
|  | Right Frontal Lobe Activation | 0.104 | 0.249 | 0.030* |
|  | Left Temporal Lobe Activation | 0.097 | 0.097 | 0.003* |
|  | Right Temporal Lobe Activation | 0.024* | 0.768 | 0.008* |
|  | VABS Adaptive Behavior Composite | 0.000* | 0.000* | 0.000* |
|  | MSEL Receptive Language | 0.000* | 0.000* | 0.000* |
|  | MSEL Expressive Language | 0.000* | 0.000* | 0.000* |
| External | ADOS Total Score | 0.193 | 0.015* | 0.003* |
|  | MSEL Early Learning Composite | 0.000* | 0.000* | 0.000* |
|  | GeoPref Social Fixation | 0.891 | 0.195 | 0.195 |

*Note.* Multiple pairwise comparisons were corrected by FDR, \* = $P < .05$ .

**Figure 2.**
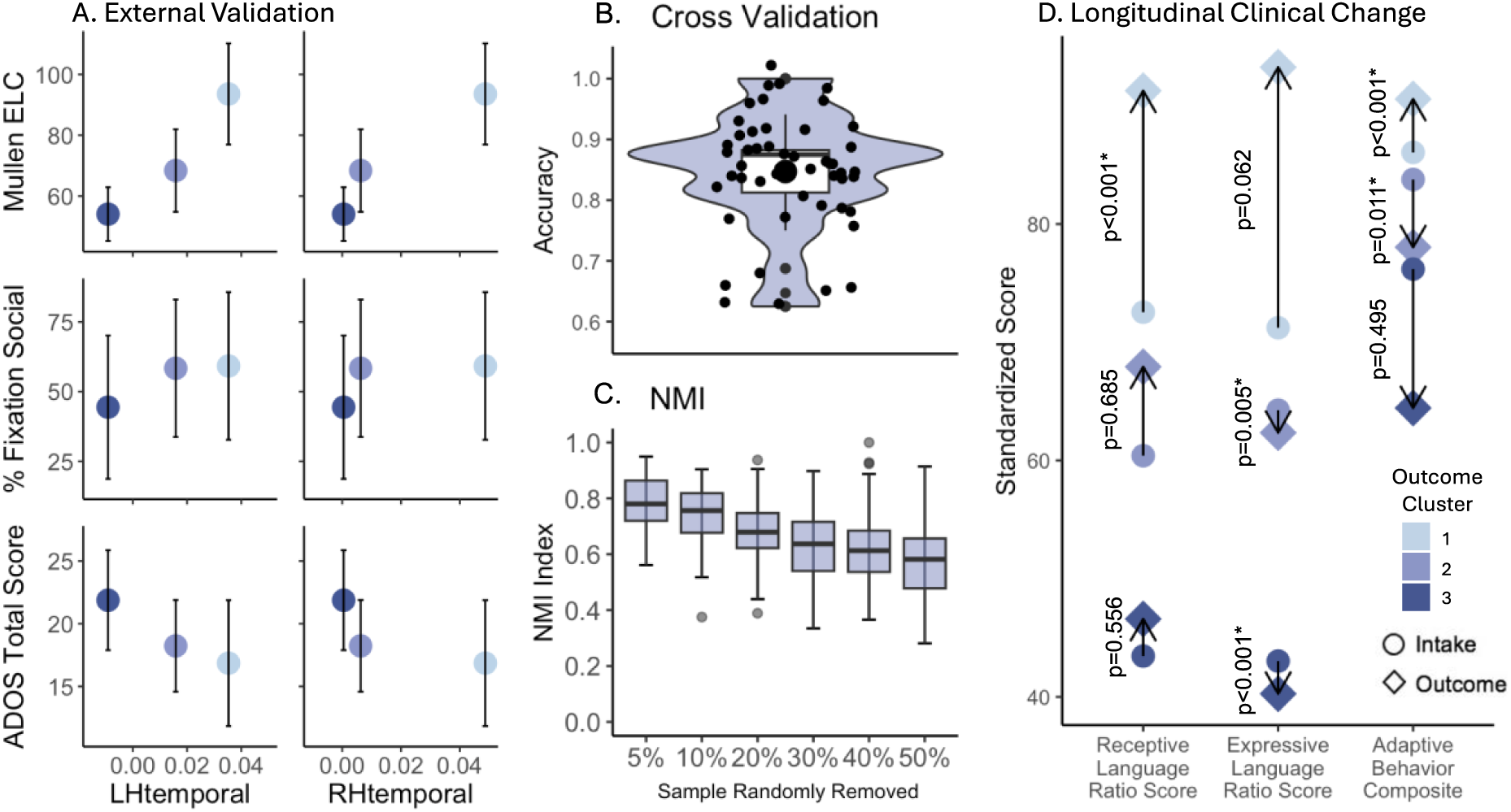
ASD Outcome SNF Validation and Longitudinal Clinical Change. (**A**) Comparing temporal lobe activation scores to clinical measures excluded from SNF clustering (IQ, social attention during eye tracking, and severity on the ADOS). See Tables 2 and 3 for statistical differences across cluster separation, all together and pairwise. (**B**) 5-fold cross validation of outcome clustering. (**C**) NMI index scores for clustering after random removal of 5% to 50% of the sample. (**D**) Longitudinal clinical scores for clusters across intake (circle) and outcome (diamond) assessments with arrows to indicate direction and magnitude of clinical change, showing ASD Outcome cluster 1 subjects have significant age-related clinical improvement, but ASD Outcome cluster 3 subjects have no clinical improvement. Significant change over time is denoted with an asterisk (p<0.05).

Third, 5-fold cross-validation resulted in a mean cluster classification of 0.85 ± 0.1SD (Figure 2B). Fourth, NMI indexing after random removal of 5% to 50% of the sample resulted in high cluster robustness even after removal of up to 30% of the total sample (i.e., leaving only 56 total subjects for clustering) (Figure 2C).

#### Clinical Change of ASD Outcome Clusters

To test for subtype-specific differences in trajectories of longitudinal clinical language and adaptive behavior abilities, we tested whether longitudinal changes between each child’s intake and outcome clinical scores (the latter of which were used in SNF and best estimated the child’s relatively ultimate phenotype) differed by cluster. Figure 2D shows significant intake-to-outcome clinical differences between the three ASD clusters. For ASD Outcome cluster 1, language scores improved substantially, while for ASD Outcome cluster 3 (the profound autism cluster) clinical scores changed little or decreased. Table 4 details statistical effects. Most importantly, this analysis revealed that patients in the mild ASD subtype improved substantially between intake and outcome assessments, but those in the profound autism subtype do not, highlighting the important clinical translational potential of multimodality subtyping.

**Table 4.** Longitudinal Clinical Change – intake to outcome change in clinical variables.

| *Cluster* | *Subscale* | *Statistic* | *P-value* |
| --- | --- | --- | --- |
| ASD Outcome 1 | VABS Adaptive Behavior Composite | 4.44<sup>a</sup> | 0.000* |
|  | MSEL Receptive Language | 5.79<sup>a</sup> | 0.000* |
|  | MSEL Expressive Language | 1.93<sup>a</sup> | 0.062 |
| ASD Outcome 2 | VABS Adaptive Behavior Composite | 354<sup>b</sup> | 0.011* |
|  | MSEL Receptive Language | 212<sup>b</sup> | 0.685 |
|  | MSEL Expressive Language | 91.5<sup>b</sup> | 0.005* |
| ASD Outcome 3 | VABS Adaptive Behavior Composite | 82<sup>b</sup> | 0.495 |
|  | MSEL Receptive Language | -0.6<sup>a</sup> | 0.556 |
|  | MSEL Expressive Language | 0.0<sup>b</sup> | 0.000* |

Note. \* = $P < .05$ , <sup>a</sup>: t-test, <sup>b</sup>: Exact Wilcoxon signed rank test

### <u>Result 2:</u> ASD subtypes in the context of non-ASD toddlers

Having delineated and validated three ASD Outcome clusters with distinct brain-clinical subtypes of ASD, including a profound autism subtype, we further investigated how these three ASD subtypes compare to typically developing (TD) and developmentally delayed (Delay) toddlers, such as language delay or global developmental delay. To do so, we expanded the cohort from the previous ASD-only analysis to include an additional 33 TD and 23 Delay subjects for an overall total of 137 toddlers. The analysis revealed three neurofunctional and clinically distinct SNF clusters, referred to here as “Mixed” clusters (Figure 3). Only 17% of higher ability ASD Outcome cluster 1 were in Mixed cluster 1 which is comprised of largely higher ability TD and Delay subjects (n=17 and n=3, respectively) (Figure 3; Table 5). The majority of ASD Outcome cluster 1 (74%) along with a subset of ASD Outcome cluster 2 (33%) were in Mixed cluster 2 (n=69), otherwise comprised of somewhat lower ability TD and Delay subjects (n=16 and n=17, respectively). Since 17% of ASD Outcome cluster 1 were in the high ability Mixed cluster 1, this suggests the importance of our Mixed cluster approach in identifying, among the best outcome ASD toddlers, the subset who may have not just a good outcome but possibly an optimal outcome given their clinical scores. Thus, a large percentage of good ability ASD toddlers clustered either with lower ability TD and Delayed toddlers or with high ability TD.

**Figure 3.**
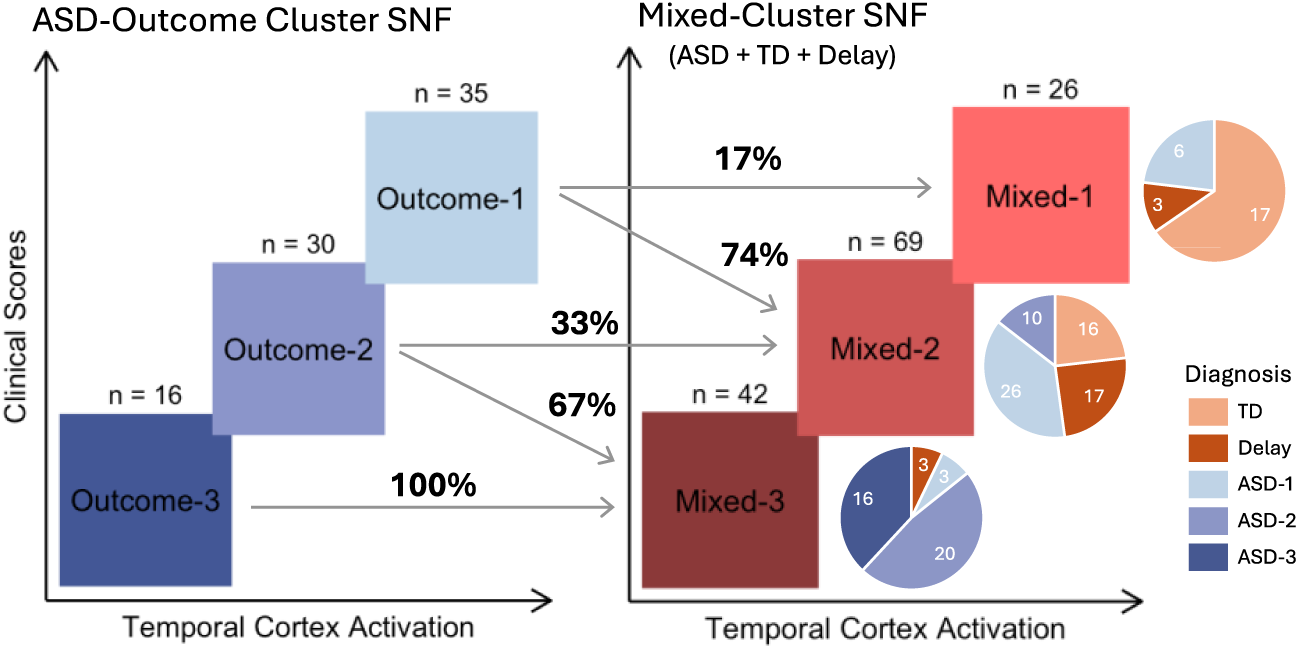
Schematic representation of mapping between clustering ASD subjects alone (blue) and clustering subjects across diagnostic categories (red). Pie charts indicate the proportion of subjects from each diagnostic category that make up each mixed cluster.

**Table 5.** Average fMRI activation in temporal cortex and clinical ability scores for each Mixed cluster and for diagnostic groups within each cluster.

|  | MSEL Receptive Language | MSEL Expressive Language | ADOS Total Score | Left Temporal Lobe Activation | Right Temporal Lobe Activation |
| --- | --- | --- | --- | --- | --- |
| **Mixed 1** | **112.67** ± 15.56 | **113.52** ± 12.93 | **5.04** ± 6.20 | **0.0432** ± 0.05 | **0.0595** ± 0.06 |
| ASD (*n*=6) | 101.49 ± 14.82 | 113.21 ± 6.09 | 15.17 ± 4.62 | 0.0081 ± 0.03 | 0.0546 ± 0.06 |
| Delay (*n*=3) | 110.89 ± 23.04 | 106.39 ± 16.96 | 3.33 ± 1.15 | 0.0403 ± 0.05 | 0.0601 ± 0.04 |
| TD (*n*=17) | 116.93 ± 13.29 | 114.89 ± 14.22 | 1.76 ± 1.68 | 0.0562 ± 0.05 | 0.0611 ± 0.06 |
| **Mixed 2** | **89.13** ± 15.24 | **87.39** ± 15.65 | **11.04** ± 7.67 | **0.0342** ± 0.05 | **0.0356** ± 0.08 |
| ASD (*n*=36) | 86.74 ± 14.83 | 85.70 ± 15.01 | 17.44 ± 3.70 | 0.0370 ± 0.05 | 0.0333 ± 0.07 |
| Delay (*n*=17) | 87.35 ± 17.71 | 84.37 ± 19.99 | 4.41 ± 4.56 | 0.0254 ± 0.06 | 0.0439 ± 0.12 |
| TD (*n*=16) | 96.39 ± 11.49 | 94.41 ± 9.44 | 3.69 ± 2.60 | 0.0372 ± 0.03 | 0.0319 ± 0.07 |
| **Mixed 3** | **56.94** ± 15.12 | **51.35** ± 17.57 | **18.79** ± 6.02 | **0.0053** ± 0.04 | **0.0062** ± 0.06 |
| ASD (*n*=39) | 57.59 ± 15.36 | 51.64 ± 18.13 | 19.69 ± 5.16 | 0.0047 ± 0.05 | 0.0096 ± 0.06 |
| Delay (*n*=3) | 48.60 ± 9.14 | 47.59 ± 8.11 | 7.00 ± 3.61 | 0.0138 ± 0.02 | -0.0370 ± 0.03 |
| TD (*n*=0) | -- | -- | -- | -- | -- |

In contrast, 100% of the profound ASD subjects from ASD Outcome cluster 3 were also in the lowest ability Mixed cluster 3. This was notably the only Mixed cluster to contain profound ASD subjects based on ASD Outcome clusters. 67% of moderate clinical ability ASD Outcome cluster 2 also fall into this lowest ability Mixed cluster 3. Mixed cluster 3 subjects (which included all profound autism subjects) had cognitive and language scores about 2 standard deviations below average and near zero brain activation in response to speech. On the other hand, Mixed cluster 1 showed average or above average clinical scores and high neural responses. Subjects in Mixed cluster 2 were in the mid to low average range across clinical and neural scores (Figure 4A, Table 5). Generally, in the Mixed clusters, activation of both the right and left temporal cortices, as well as the right frontal cortex, was significantly correlated with MSEL receptive and expressive language scores (Supplementary Table 2). There was no correlation between the left frontal cortex and language scores.

**Figure 4.**
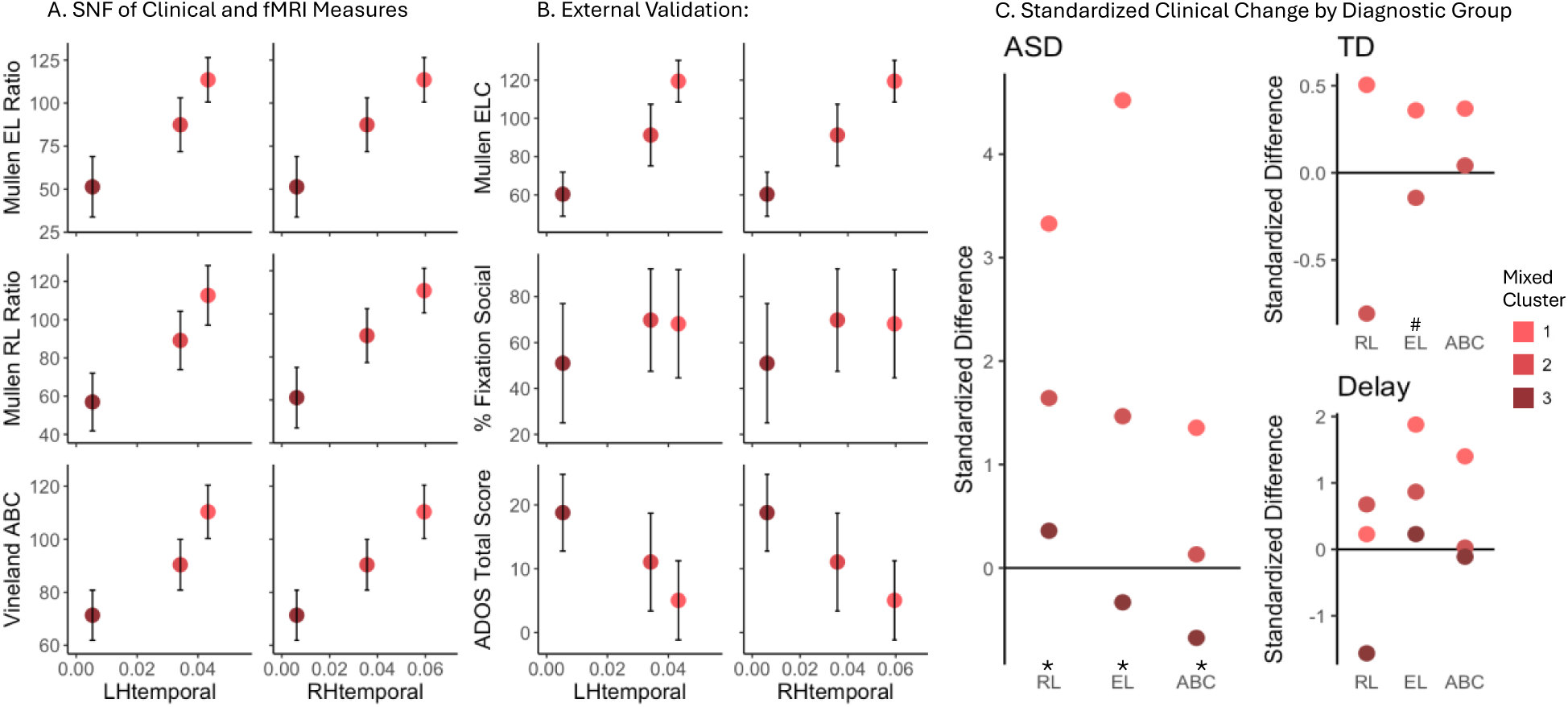
Mixed SNF Clustering. Observed relationship between temporal brain activation and clinical scores (**A**) in Mixed-Cluster SNF and (**B**) external validation using clinical scores excluded from Mixed-Cluster SNF (for specific details, see Figure 1). (**C**) Longitudinal clinical scores for each diagnostic group in each cluster, each separated by Mixed cluster assignment. Zero standardized difference indicates no group level change across visits. Statistical significance of cluster separation based on each variable through nonparametric ANCOVAs are denoted with an asterisk (p<0.05) or a pound sign (p<0.1), detailed in Supplemental Tables 2 and 3.

#### Validation of Mixed Clusters

As for the ASD Outcome clusters, we next validated SNF Mixed clusters in multiple ways by testing cluster separation, external variables, 5-fold cross validation and robustness. Both SNF variables and external variables contributed individually to significant separation of the three Mixed clusters, except for the left frontal lobe activation (see Tables 6 and 7). The ANOVA and pairwise comparisons showed clear separation between clusters with respect to SNF clinical variables as well as two external clinical variables (ADOS total score and MSEL ELC). Regarding the activation of the right and left temporal cortices and the external eye tracking score, the Mixed cluster 3 was distinctly separated from both the medium and high ability Mixed clusters 2 and 1. For right frontal cortex activation, only Mixed cluster 3 was significantly different from Mixed cluster 2. External clinical and social attention scores are shown in relation to temporal cortex fMRI activation in Figure 4B. 5-fold cross-validation shows mean cluster classification is 0.79 ± 0.1SD (see Supplemental Figure 1). NMI indexing after random removal of 5% to 50% of the sample showed robustness even after removal of up to 40% of the total sample (i.e., leaving only 82 total subjects for clustering) (Supplemental Figure 1).

**Table 6.** Mixed Cluster Separation Validation and External Validation in Mixed SNF variables.

|  | *Variable* | *Effect* | *F-ratio* | *DFn* | *DFd* | *GES* | *P-value* |
| --- | --- | --- | --- | --- | --- | --- | --- |
| SNF | Left Frontal Lobe Activation | Clusters | 2.03 | 2 | 134 | - | 0.135 |
|  | Right Frontal Lobe Activation | Clusters | 3.28 | 2 | 134 | 0.05 | 0.041* |
|  | Left Temporal Lobe Activation | Clusters | 6.87 | 2 | 134 | 0.09 | 0.001* |
|  | Right Temporal Lobe Activation | Clusters | 4.7 | 2 | 134 | 0.07 | 0.011* |
|  | VABS Adaptive Behavior Composite | Clusters | 135.04 | 2 | 134 | 0.67 | 0.000* |
|  | MSEL Receptive Language | Clusters | 115.75 | 2 | 134 | 0.63 | 0.000* |
|  | MSEL Expressive Language | Clusters | 134.42 | 2 | 134 | 0.67 | 0.000* |
| External | ADOS Total Score | Clusters | 33.73 | 2 | 134 | 0.34 | 0.000* |
|  | MSEL Early Learning Composite | Clusters | 150.01 | 2 | 134 | 0.69 | 0.000* |
|  | GeoPref Social Fixation | Clusters | 8.07 | 2 | 123 | 0.12 | 0.001* |

*Note.* \* = $P < .05$ , *DFn* = Degrees of freedom for numerator, *DFd* = Degrees of freedom for denominator, *GES* = Generalized Eta Squared (effect size).

**Table 7.** Separation Validation. Mixed Cluster *<u>pairwise</u>* comparisons in Mixed SNF variables and external variables.

|  | *Variable* | *P-values* |  |  |
| --- | --- | --- | --- | --- |
|  | Mixed Cluster | 1 - 2 | 2 - 3 | 1 - 3 |
| SNF | Left Frontal Lobe Activation | 0.165 | 0.165 | 0.746 |
|  | Right Frontal Lobe Activation | 0.432 | 0.022* | 0.142 |
|  | Left Temporal Lobe Activation | 0.409 | 0.003* | 0.003* |
|  | Right Temporal Lobe Activation | 0.144 | 0.015* | 0.001* |
|  | VABS Adaptive Behavior Composite | 0.000* | 0.000* | 0.000* |
|  | MSEL Receptive Language | 0.000* | 0.000* | 0.000* |
|  | MSEL Expressive Language | 0.000* | 0.000* | 0.000* |
| External | ADOS Total Score | 0.001* | 0.000* | 0.000* |
|  | MSEL Early Learning Composite | 0.000* | 0.000* | 0.000* |
|  | GeoPref Social Fixation | 0.815 | 0.001* | 0.017* |

*Note.* Multiple pairwise comparisons were corrected by FDR, \* = $P < .05$ .

#### Clinical Change in Different Diagnostic Groups in Mixed Clusters

Next, we tested whether changes between each child’s intake and outcome clinical scores (the latter of which were used in SNF, and best estimated the child’s stable preschool outcome phenotype) differed by cluster. Subjects in Mixed cluster 1 showed significant improvement in language scores between intake and outcome ages, while subjects in Mixed cluster 3 showed nearly null change or a decrease. Subjects in Mixed cluster 2 showed slight improvement. (Supplemental Figure 1, see statistical results in Supplemental Tables 3-4). When intake-to-outcome clinical change scores are considered separately for ASD, TD and Delayed subjects within each Mixed cluster (Figure 4C, Supplemental Tables 5-6), we find that ASD subjects in Mixed cluster 1 show significant clinical improvement in the range of 2-4 standard deviations from their initial scores. ASD subjects in Mixed cluster 2 showed improvement to a lesser degree, around 0 to 2 standard deviations. ASD subjects in Mixed cluster 3 showed insignificant changes, including slight decline in some clinical measures. TD subjects in Mixed cluster 1 showed greater improvements than those in Mixed cluster 2, however the TD subjects were overall more stable across visits and only showed improvement or decline less than a standard deviation from their baseline scores. Delay subjects in Mixed cluster 1 showed improvement in most areas achieving neurotypical scores, and those in Mixed cluster 2 showed minimal change with trending improvement. The few Delay subjects in Mixed cluster 3 showed some clinical measures declined and others neither improved nor declined.

### <u>Result 3:</u> Independent replication dataset further validates Mixed Cluster results

To assess the reproducibility of our Mixed cluster findings, we repeated SNF with a completely independent cohort of ASD, TD, and Delay subjects from our laboratory’s previous work (Xiao et. al., Nature Human Behaviour^6^), referred to here as “NHB” clusters. Despite an overall sample of only 42 subjects with all required measurements, the replication clustering resulted in three clusters with similar diagnostic distributions as was found in Mixed clustering (Figure 5A). Both NHB and Mixed cluster 3 were 87-93% ASD and 7-13% Delay, with no TD subjects. In contrast, NHB and Mixed cluster 1 were 62-65% TD with only 23% ASD subjects. NHB and Mixed cluster 2 were both equally ASD (50-52%) and non-ASD, with a slightly higher level of Delay (25-36%) than TD (14-23%) subjects. These clusters also depicted brain-behavior subtypes with a similar pattern of features, including NHB cluster 1 (n=13) with high clinical scores and high brain activation, NHB cluster 3 (n=15) with low clinical scores and low brain activation, and NHB cluster 2 (n=14) that falls in the average range both clinically and neurologically (Figure 5C, Table 8).

**Figure 5.**
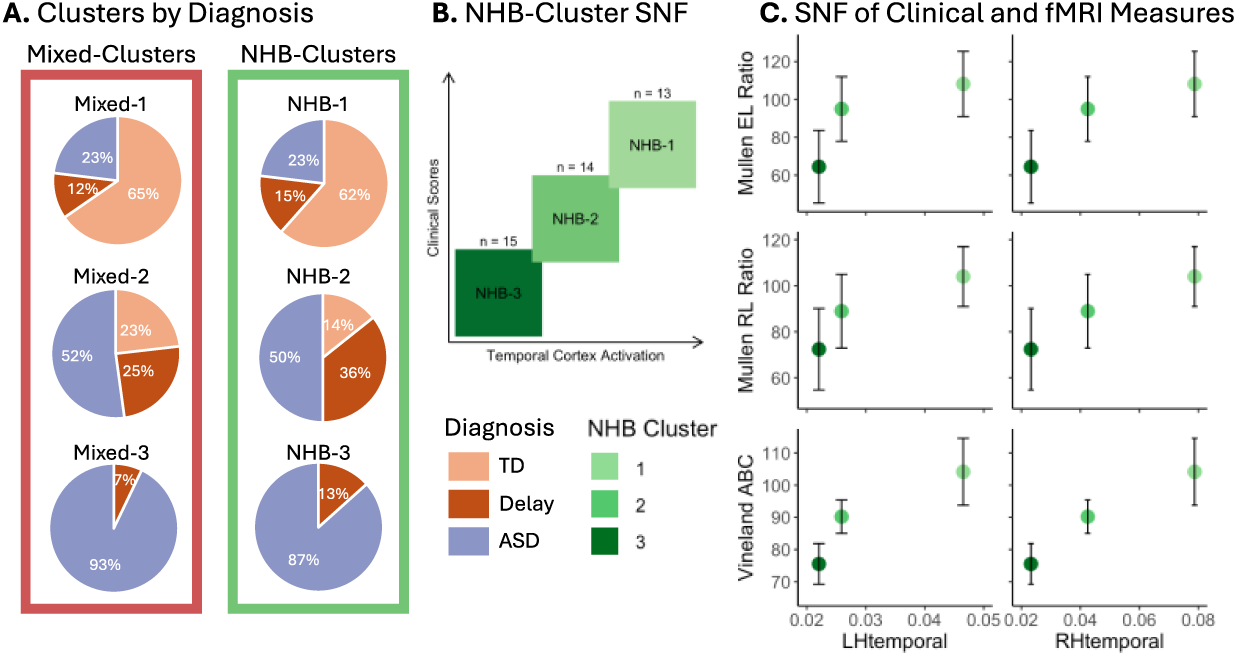
Independent SNF Cluster Replication. (**A**) Proportion of subjects with each diagnosis assigned to each cluster in mixed clustering (red outline) and NHB clustering (green outline). (**B**) Schematic representation of independent cohort clustering. (**C**) Observed relationship between temporal brain activation and clinical scores used in SNF for NHB clusters (for specific details, see Figure 1B).

**Table 8.** Average fMRI activation in temporal cortex and clinical ability scores for each NHB cluster and for diagnostic groups within each cluster.

|  | MSEL Receptive<br>Language | MSEL Expressive<br>Language | ADOS<br>Total Score | Left Temporal<br>Lobe Activation | Right Temporal<br>Lobe Activation |
| --- | --- | --- | --- | --- | --- |
| **NHB 1** | **103.99** ± 13.02 | **108.20** ± 17.19 | **7.31** ± 6.75 | **0.0465** ± 0.03 | **0.0787** ± 0.06 |
| *ASD* | 93.43 ± 8.96 | 115.09 ± 6.43 | 18.67 ± 4.04 | 0.0307 ± 0.02 | 0.0364 ± 0.06 |
| *Delay* | 106.90 ± 8.11 | 101.52 ± 3.71 | 4.50 ± 0.71 | 0.0479 ± 0.05 | 0.1181 ± 0.06 |
| *TD* | 107.22 ± 14.01 | 107.28 ± 21.42 | 3.75 ± 1.16 | 0.0521 ± 0.03 | 0.0848 ± 0.05 |
| **NHB 2** | **88.94** ± 16.03 | **94.99** ± 17.04 | **10.93** ± 8.21 | **0.0259** ± 0.03 | **0.0425** ± 0.04 |
| *ASD* | 89.54 ± 18.71 | 101.94 ± 17.53 | 18.43 ± 3.46 | 0.0163 ± 0.01 | 0.0310 ± 0.01 |
| *Delay* | 85.69 ± 16.12 | 80.91 ± 7.28 | 3.80 ± 1.92 | 0.0294 ± 0.03 | 0.0456 ± 0.04 |
| *TD* | 94.99 ± 8.51 | 105.86 ± 12.44 | 2.50 ± 0.71 | 0.0509 ± 0.06 | 0.0748 ± 0.11 |
| **NHB 3** | **72.39** ± 17.72 | **64.47** ± 19.18 | **16.40** ± 6.65 | **0.0221** ± 0.02 | **0.0232** ± 0.03 |
| *ASD* | 73.84 ± 17.99 | 64.60 ± 20.51 | 18.31 ± 4.70 | 0.0207 ± 0.02 | 0.0212 ± 0.03 |
| *Delay* | 62.93 ± 17.49 | 63.63 ± 10.12 | 4.00 ± 0.00 | 0.0311 ± 0.04 | 0.0363 ± 0.04 |
| *TD* | -- | -- | -- | -- | -- |

### <u>Result 4:</u> Subtyping ASD at toddler-ages yields less translationally reliable clusters: Temporal instability

Pierce et al (2019)^32^ reported that once a toddler is diagnosed with ASD, there is high stability of that ASD diagnosis at follow-up ages. On the other hand, Pierce et al also found that 24% of 12 to 24 month olds diagnosed with ASD later, at ages 2 to 4 years, had clinical characteristics of typical or mildly delayed toddlers at their *earlier ages* of 12 to 24 months. Thus, some later-age diagnosed ASD toddlers showed pronounced age-related shifts in phenotype. Longitudinal validation analyses of the three ASD Outcome clusters above showed striking differences between clusters in longitudinal change scores from intake ages to outcome ages, with ASD Outcome 1 patients having substantial improvement in clinical scores as opposed to profound ASD Outcome 3 remaining low across intake to outcome. Thus, the Pierce et al study and our present evidence warrant the hypothesis that using some types of early age clinical data to cluster toddlers with SNF may lead to results poorly aligned with later SNF outcome clusters, which utilize scores at ages 2 – 4 years as a “best estimate” of a child’s stable clinical phenotype. To test this hypothesis in an unbiased manner separately from the Pierce et al study, SNF was repeated for the same 81 ASD subjects using clinical scores *<u>at intake</u>* rather than outcome to test if identical unsupervised clustering based on a younger timepoint would be predictive of outcome subtype. The resulting three intake clusters failed to identify a clear relationship between neural activation and clinical ability, and cluster membership at Intake poorly matched ASD Outcome cluster membership. This early-age cluster instability is also indicated by a Jaccard index of only 31% when Intake and Outcome clusters are compared and is illustrated in Figure 6, showing that most ASD toddlers in each Intake cluster switch to different Outcome clusters. For instance, only 40% of Intake cluster 3 stay in Outcome cluster 3. Thus, some clinical scores at very early ages are in flux and are neither stable nor reliable clustering indicators of whether a child will improve or decline across development (Supplemental Figure 2, Supplemental Table 7-9).

**Figure 6.**
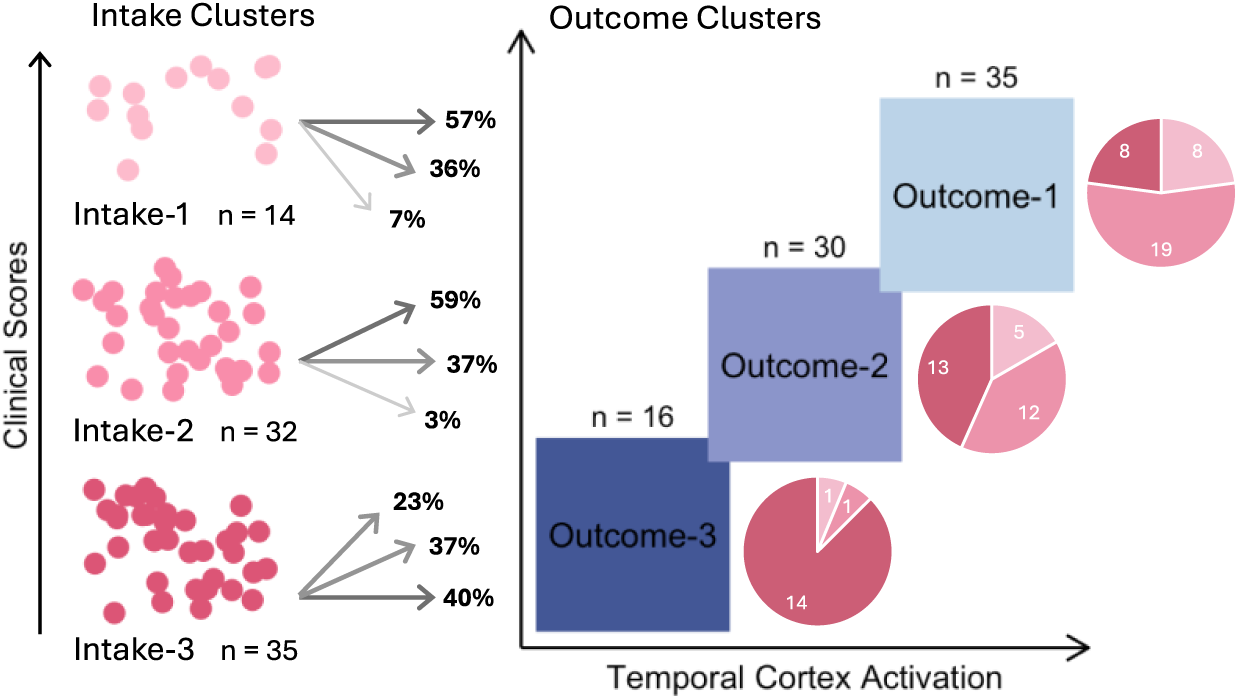
Schematic representation of SNF clustering using Intake scores (pink), and how individual subjects map to SNF clusters based on Outcome scores (blue).

## DISCUSSION

Once the critical leap from treatment based purely on an operant model (e.g., Discrete Trial Training) to more naturalistic behavioral interventions occurred^33^, interventions for children with ASD have not changed substantially at a conceptual level in the past 30 years. Some children with ASD thrive in response interventions, and some do not; yet it is not predictable before treatment which children will do well and why, particularly at the early ages of 1 to 3 years when treatment has the potential to be most beneficial. As one study concluded, it is only possible to know which child may benefit until <u>after</u> one sees treatment outcome^34^. Precision medicine efforts to find rigorously validated ASD social neural subtypes currently do not exist, in part due to the field’s limited understanding of the biological drivers of early social heterogeneity. Of 25 clinical clustering studies of ASD toddlers^3^, none incorporated both neurobiological measures relevant to social communication symptoms and clinical scores into objective data-driven clustering, and so, none have provided insight on what might be the neural bases of social subtypes.

Here, we take advantage of current deep knowledge about the role of temporal cortex in typical social brain^10,11^ and its anatomical^15–17^, functional^4,6,24,28^ and gene expression^18^ differences in ASD by incorporating measures of temporal cortex activation by social speech with clinical scores in a within-ASD child unsupervised SNF clustering design. Additionally, the clustering results were rigorously validated using this social fMRI activation and deep phenotyping in a large sample of ASD and non-ASD toddlers. Results identified three ASD Outcome social neural-clinical subtypes with distinctly different temporal cortex social activation and clinical developmental trajectories. Subtypes were thoroughly validated, something missing from previous ASD studies, and clinically important developmental trajectories quantitatively assessed. Each ASD Outcome subtype had a different characteristic level of social and language ability and early-age developmental trajectory.

As compared to other ASD subtypes, toddlers in the “high” ASD Outcome subtype had the most robust superior temporal cortical activation by social language, the highest social and language abilities, and the greatest longitudinal clinical improvement. In contrasting subtype, toddlers in the “low” ASD subtype had the opposite phenotype, namely, weak temporal cortex social speech activation, very low social and language abilities, and persistently low clinical abilities across age. This clinical profile is consistent with profound autism^1^. As such, the SNF-derived high and low ASD Outcome subtypes were not simply quantitatively different from each other, but rather were categorically opposite clinical and neurofunctional types of ASD with one showing strong age-related improvement and the other persistently low clinical ability and severe social symptoms indicative of profound autism.

There are three principal implications of these results and one major new avenue of research opened. First, the “spectrum” of ASD heterogeneity is not truly a continuous spectrum from the neurobiological and clinical perspectives. The profound autism subtype is arguably the neurobiological opposite of the high ability and optimal outcome ASD subtype. Whereas profoundly affected toddlers have notably weak and in some cases absent neural response to social affective communications in temporal cortex (a key hub of social information processing), the high ability subtype has near neurotypical and in some cases neurotypical neural responses in right temporal cortex. Neural functional organization in profound and high ASD appear to be different from each other, a difference that highlights the need to conduct experiments specifically designed to uncover their neural organizational differences from each other, not just from neurotypical temporal cortex neural structure and function.

Moreover, profound autism is definitively the clinical opposite of the high ability and optimal outcome ASD subtype. Whereas profoundly affected toddlers have persistent and severely low language, social and cognitive abilities (in some cases even declining further with age), the high ability subtype often shows improving, even substantially improving, language and cognitive abilities; yet, patients in this subtype have minimally changing social symptoms despite near neurotypical social neural responses in temporal cortex at early ages and improving language across ages.

A second implication is that neurobiological and clinical subtype differences highlight the need to develop subtype-specific treatments, particularly for the profound subtype. It is unlikely that the same behavioral or pharmaceutical treatment of core autism symptoms will have the same beneficial effect on both profound and mild autism. The disappointing lack of significant advances in treatment strategies in the past 30 years could be due to the uncontrolled admixture of these opposite types of ASD. For example, one recent intervention meta-analysis noted: “When effect estimation was limited to RCT designs and to outcomes for which there was no risk of detection bias, no intervention types showed significant effects on any outcome.”^35^ Results from the overwhelming majority of treatment studies in that meta-analysis and elsewhere report outcomes only at the group (not subtype) level. The proportion of each subtype in any given sample likely explains why some studies find better or poorer treatment effects.

Thus, a third implication is that treatment studies with an undetermined mix of subtypes could fail or succeed based on how many patients from each subtype are included in the mix. The only path forward is by starting with knowing what subtype a toddler is and addressing each subtype separately. Treatment studies –whether behavioral or pharmaceutical--that persist in ignoring a priori subtype differences may be persisting in a failed approach to ASD treatment and should be discouraged.

The new avenue of research comes from two new ASD studies, a brain cortical organoid (BCO) study of an embryogenesis model of ASD^36^ and a toddler-age blood gene expression study (Zahiri, in progress). BCOs derived from profound autism toddlers have extreme and accelerated embryonic overgrowth and accelerated neurogenesis. These patients have substantial cortical growth differences in temporal cortex as well as other social, language, face, and sensory processing regions. Embryonic BCO size predicted the ASD social symptom severity. Thus, even at the early age of embryogenesis in these models, profound ASD neurobiology differs from mild ASD neurobiology. At toddler ages, profound ASD toddlers – but not mild ASD – have blood gene expression dysregulation of five pathways that govern embryonic cell proliferation, neurogenesis, and growth, again highlighting the important different neurobiology of profound autism as compared with mild ASD. This evidence suggests that ASD treatment research that incorporates subtyping and studying profound and high ASD separately can be properly interpreted.

To contextualize these three ASD Outcome subtypes within the broader non-ASD toddler population, we mapped and validated them with comparable data from a range of typical and developmental delayed subjects again using data driven SNF. We found that even for non-ASD groups, diagnostic category was not exclusively indicative of cluster assignment. TD and delayed groups displayed heterogeneity just as did the ASD group, each splitting into two or three subtypes: high and somewhat lower ability TD subtypes with differing temporal social activation; delayed subtypes with rapid improvement and those without. The three ASD Outcome subtypes persist, even among this range of non-ASD toddlers. 74% of those in the high ASD Outcome subtype overlapped with typical toddlers albeit with lower ability, potentially indicating a relatively good outcome long term. Importantly, overlapping with *high ability typical toddlers* were 17% of the high ability ASD Outcome subtype; these high ability ASD toddlers may eventually prove to be “optimal” outcome ASD individuals, a previously clinically described subgroup of 5-10% of the ASD population^37,38^.

At the opposite end of optimal ASD outcome, is profound autism estimated in the literature to be between 20% and 30% of the ASD population^2,39^. Here, 20% of our total ASD sample had a profound clinical phenotype. Profound autism subjects in the present study had negligible social language activation of temporal social cortex. Thus, data driven multimodal SNF performed both within ASD alone and among a range of non-ASD development provided precision information at early preschool ages about which ASD child has the potential to achieve the best outcomes and which may struggle with profound autism across a lifetime. Knowledge of an ASD child’s subtype is of the utmost importance for parents, professionals and service providers^40^, and would help each child according to their individual needs. Currently such precision knowledge does not exist in clinical practice for ASD^41,42^ and affected children and families suffer as a result. Yet, obtaining that subtype information by 3 years of age is possible based the present SNF brain-clinical data.

Although our earliest age-point yielded clusters less clinically reliable as predictors when considered independently, this intake clustering could still provide meaningful insight to a toddler’s short-term trajectory. For instance, toddlers in the high ASD Intake cluster are very *<u>unlikely</u>* to end up in the low ASD Outcome cluster and instead are more likely to show a good chance of improvement. Conversely, about half the toddlers in the low ASD Intake cluster map to the low ASD Outcome cluster; however, many others ended up among higher ability ASD subtypes, suggesting that at the earliest ages, some delayed clinical abilities might not necessarily be a precursor of a negative outcome. This group is similar to so-called “late-talkers”^20^ and discrimination of toddlers showing these targetable delays from those showing early signs of severe ASD symptomology warrants further investigation. It can be hypothesized that many toddlers in this cluster would benefit most from very early treatments, and perhaps the failure to detect this subtype early on contributes to the lack of improvement at later ages.

In conclusion, we think the three social neural-clinical ASD subtypes will be found to have different underlying molecular pathobiology and thus, will benefit from different biological treatments. Research to develop clinical translation of multimodal SNF patient subtyping may one day lead to precision methods for determining which subtype an individual ASD toddler patient belongs to. Objective data-driven separation has the potential^5,43^ to improve early-age subtype-specific detection, treatment, understanding of mechanisms via ASD iPSC models, and ultimately causal factors. Lastly, it can be anticipated that with larger sample sizes and with the addition of molecular data and other imaging, behavioral, and clinical data types, these 3 neural-clinical subtypes are likely to be more precisely and accurately characterized at still earlier ages and perhaps subdivided into additional subtypes. That would advance clinical care, treatment development and selection, and discovery of underlying biological causes and processes. At this stage in our ASD knowledge, subtypes should not be considered fixed or immutable, but rather a powerful step towards advancing knowledge of the underlying neural biological condition and increasing clinical benefit for patients with ASD.

## Data Availability

All data produced in the present study are available upon reasonable request to the authors.

