## Supplemental Figures & Tables for "Multimodality Integration of Neural Social Activation and Social and Language Scores Reveals Three Replicable Profound and Milder Autism Subtypes With Divergent Clinical Outcomes"

*Senior Biostatistician First Author

### Co-Senior Author

#### Senior Author

**Correspondence to:

Vani Taluja
Autism Center of Excellence, Department of Neurosciences, University of California, San Diego, La Jolla, CA, 92037, USA


Eric Courchesne
Autism Center of Excellence, Department of Neurosciences, University of California, San Diego, La Jolla, CA, 92037, USA


#### Supplementary Information

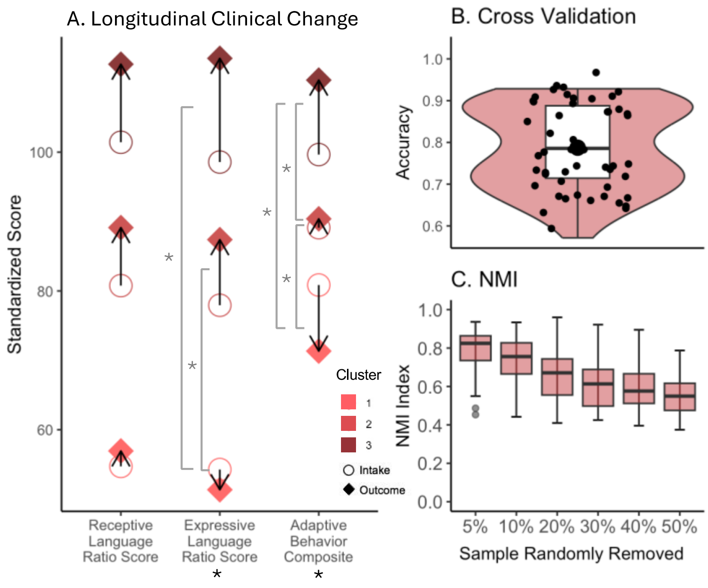


**Supplemental Figure 1**. Validations of Mixed clustering results. (**A**) Change in clinical scores across intake (open circle) and outcome (filled diamond) assessments with arrows to indicate direction and magnitude of change. Significant pairwise (gray) and all cluster (black) separation based on change score is denoted with an asterisk ($p<0.05$), detailed in Supplemental Tables 4 and 5. (**B**) 5-fold cross validation of mixed clustering. (**C**) NMI index scores for clustering after random removal of 5% to 50% of the sample.

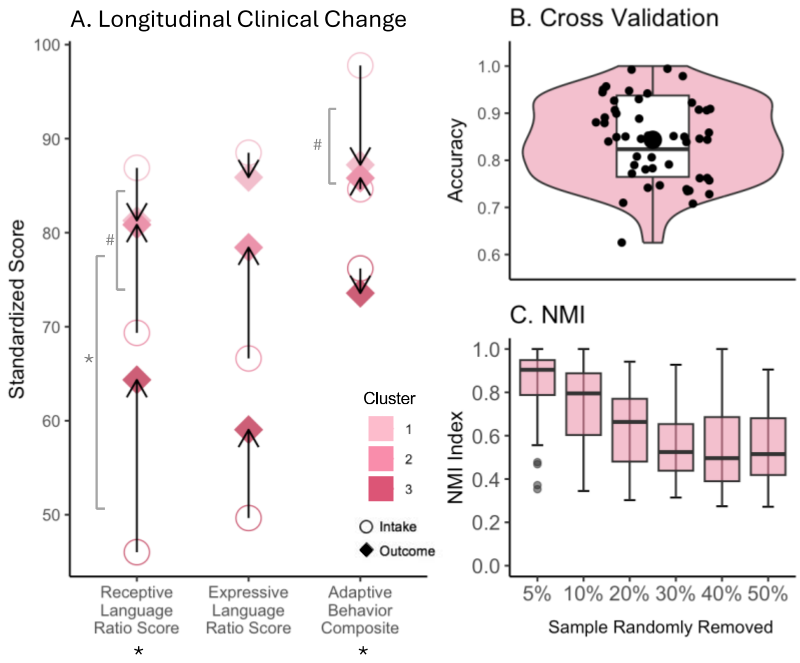


**Supplemental Figure 2**. Validations of Intake clustering results. (**A**) change in clinical scores across intake (open circle) and outcome (filled diamond) assessments with arrows to indicate direction and magnitude of change. Significant pairwise (gray) and all cluster (black) separation based on change score is denoted with an asterisk ($p<0.05$) or a pound sign ($p<0.1$), detailed in Supplemental Tables 6 and 7. (**B**) 5-fold cross validation of intake clustering. (**C**) NMI index scores for clustering after random removal of 5% to 50% of the sample.

**Supplemental Table 1.** Subject demographics and clinical characteristics

| | ASD (n=81) | TD (n=33) | Delay (n=23) | Total (n=137) |
|---|---|---|---|---|
| *Demographics* | | | | |
| Sex (M/F) | 65 / 16 | 24 / 9 | 18 / 5 | 107 / 30 |
| Age at clinical Intake (months) | 22.62 (6.23) | 15.13 (3.78) | 17.46 (6.29) | 19.95 (6.59) |
| Age at MRI (months) | 29.75 (8.30) | 25.97 (9.44) | 25.30 (8.65) | 28.09 (8.81) |
| Age at clinical Outcome (months) | 37.79 (6.18) | 31.20 (4.29) | 37.19 (9.25) | 36.10 (6.96) |
| *Intake Clinical Scores* | | | | |
| VABS Adaptive Behavior Composite | 83.26 (10.49) | 101.12 (9.91) | 89.35 (10.14) | 88.58 (12.63) |
| MSEL Receptive Language | 62.30 (21.17) | 108.29 (33.07) | 82.11 (23.72) | 76.70 (31.35) |
| MSEL Expressive Language | 63.07 (20.97) | 103.81 (18.55) | 73.29 (26.12) | 74.60 (27.14) |
| MSEL Early Learning Composite | 74.56 (16.64) | 107.12 (14.17) | 85.26 (20.60) | 84.20 (21.49) |
| ADOS Total Score | 18.74 (5.83) | 3.30 (3.91) | 9.78 (7.47) | 13.52 (8.75) |
| GeoPref Social Fixation | 56.20 (25.90) | 75.12 (19.53) | 71.45 (21.06) | 63.67 (25.03) |
| *Outcome Clinical Scores* | | | | |
| VABS Adaptive Behavior Composite | 80.77 (14.03) | 104.27 (11.31) | 92.17 (14.00) | 88.34 (16.61) |
| MSEL Receptive Language | 73.80 (21.98) | 106.97 (16.09) | 85.37 (23.67) | 83.73 (25.02) |
| MSEL Expressive Language | 71.34 (25.89) | 104.96 (15.84) | 82.44 (23.86) | 81.30 (27.21) |
| MSEL Early Learning Composite | 76.44 (21.26) | 111.64 (13.24) | 89.61 (24.78) | 87.13 (24.92) |
| ADOS Total Score | 18.36 (4.69) | 2.70 (2.35) | 4.61 (4.18) | 12.28 (8.44) |
| GeoPref Social Fixation | 56.20 (25.90) | 75.12 (19.53) | 71.45 (21.06) | 63.67 (25.03) |

*Note.* All are shown as: mean (standard deviation).

**Supplemental Table 2.** Correlation of language abilities and brain activation

| | | ASD Outcome | | Mixed | |
|---|---|---|---|---|---|
| *Brain Activation* | *MSEL Language Subscale* | *Pearson's r* | *P-value* | *Pearson's r* | *P-value* |
| Left Frontal Lobe | Receptive | 0.08 | 0.488 | 0.04 | 0.634 |
| | Expressive | -0.04 | 0.733 | -0.01 | 0.912 |
| Right Frontal Lobe | Receptive | 0.33 | 0.003* | 0.19 | 0.023* |
| | Expressive | 0.26 | 0.017* | 0.2 | 0.017* |
| Left Temporal Lobe | Receptive | 0.27 | 0.016* | 0.26 | 0.002* |
| | Expressive | 0.21 | 0.066 | 0.27 | 0.002* |
| Right Temporal Lobe | Receptive | 0.26 | 0.021* | 0.24 | 0.004* |
| | Expressive | 0.24 | 0.028* | 0.26 | 0.003* |

*Note.* * = $P < .05$.

**Supplemental Table 3.** Mixed cluster separation in clinical change scores

| *Variable* | *Effect* | *F-ratio* | *DFn* | *DFd* | *GES* | *P-value* |
|---|---|---|---|---|---|---|
| VABS Adaptive Behavior Composite | Clusters | 30.67 | 2 | 134 | 0.31 | 0.000* |
| MSEL Receptive Language | Clusters | 1.19 | 2 | 134 | - | 0.307 |
| MSEL Expressive Language | Clusters | 6.42 | 2 | 134 | 0.09 | 0.002* |

*Note*. * = *P* < .05, *DFn* = Degrees of freedom for numerator, *DFd* = Degrees of freedom for denominator, *GES* = Generalized Eta Squared (effect size).

**Supplemental Table 4.** Mixed cluster pairwise comparisons in clinical change scores

| *Variable* | *P-values* | | |
|---|---|---|---|
| Mixed Cluster | 1 - 2 | 2 - 3 | 1 - 3 |
| VABS Adaptive Behavior Composite | 0.002* | 0.000* | 0.000* |
| MSEL Receptive Language | 0.640 | 0.246 | 0.640 |
| MSEL Expressive Language | 0.376 | 0.008* | 0.008* |

*Note*. Multiple pairwise comparisons were corrected by FDR, * = *P* < .05.

**Supplemental Table 5.** Nonparametric ANCOVA for corrected change scores in Mixed SNF by diagnostic group

| | | ASD | | TD | |
|---|---|---|---|---|---|
| *Variable* | *Effect* | *h Statistic* | *P-value* | *h Statistic* | *P-value* |
| VABS Adaptive Behavior Composite | Clusters | 1.09 | 0.004* | 1.00 | 0.317 |
| MSEL Receptive Language | Clusters | 1.10 | 0.002* | 1.02 | 0.821 |
| MSEL Expressive Language | Clusters | 1.10 | 0.001* | 1.02 | 0.052 |

*Note*. * = *P* < .05

**Supplemental Table 6.** Nonparametric ANCOVA pairwise comparisons for corrected change scores in Mixed SNF- ASD

| *Variable* | *P-values* | | |
|---|---|---|---|
| Mixed Cluster | 1 - 2 | 2 - 3 | 1 - 3 |
| VABS Adaptive Behavior Composite | 0.015* | 0.043* | 0.011* |
| MSEL Receptive Language | 0.011* | 0.011* | 0.011* |
| MSEL Expressive Language | 0.009* | 0.018* | 0.009* |

*Note*. Multiple pairwise comparisons were corrected by FDR, * = *P* < .05.

**Supplemental Table 7.** Average fMRI activation in temporal cortex and clinical ability scores for each Intake cluster.

| | MSEL Receptive Language | MSEL Expressive Language | ADOS Total Score | Left Temporal Lobe Activation | Right Temporal Lobe Activation |
|---|---|---|---|---|---|
| Intake 1 | 81.29 ± 22.00 | 85.88 ± 19.12 | 18.14 ± 6.97 | 0.0102 ± 0.06 | 0.0006 ± 0.06 |
| Intake 2 | 80.85 ± 12.83 | 78.43 ± 12.87 | 17.75 ± 5.53 | 0.0326 ± 0.03 | 0.0435 ± 0.06 |
| Intake 3 | 64.35 ± 12.28 | 59.05 ± 16.74 | 19.00 ± 2.94 | 0.0108 ± 0.05 | 0.0142 ± 0.07 |

**Supplemental Table 8.** Intake cluster separation in clinical change scores

| *Variable* | *Effect* | *F-ratio* | *DFn* | *DFd* | *GES* | *P-value* |
|---|---|---|---|---|---|---|
| VABS Adaptive Behavior Composite | Clusters | 3.86 | 2 | 78 | 0.09 | 0.025* |
| MSEL Receptive Language | Clusters | 6.15 | 2 | 78 | 0.14 | 0.003* |
| MSEL Expressive Language | Clusters | 2.03 | 2 | 78 | - | 0.138 |

*Note*. * = *P* < .05, *DFn* = Degrees of freedom for numerator, *DFd* = Degrees of freedom for denominator, *GES* = Generalized Eta Squared (effect size).

**Supplemental Table 9.** Intake cluster pairwise comparisons in clinical change scores

| *Variable* | *P-values* | | |
|---|---|---|---|
| Intake Cluster | 1 - 2 | 2 - 3 | 1 - 3 |
| VABS Adaptive Behavior Composite | 0.066 | 0.235 | 0.133 |
| MSEL Receptive Language | 0.082 | 0.175 | 0.024* |
| MSEL Expressive Language | 0.115 | 0.422 | 0.115 |

*Note*. Multiple pairwise comparisons were corrected by FDR, * = *P* < .05.
